# Depth-Sensitive Cerebral Blood Flow and Low-Frequency Oscillations for Consciousness Assessment Using Time-Domain Diffuse Correlation Spectroscopy

**DOI:** 10.1101/2025.08.31.25334647

**Authors:** Sahar Sabaghian, Chien-Sing Poon, Carsi Kim, Christopher H Moore, Irfaan Dar, Timothy M. Rambo, Aaron J. Miller, Sujith Swarna, Noah Lubin, Sima Mofakkam, Charles Mikell, Jingxing Wang, Brandon Foreman, Ulas Sunar

## Abstract

This study evaluates the feasibility of depth-sensitive bedside monitoring of cerebral blood flow (CBF) and low-frequency oscillations (LFOs) using time-domain diffuse correlation spectroscopy (TD-DCS) in healthy controls and patients with disorders of consciousness (DOC). A 1064 nm TD-DCS system equipped with superconducting nanowire single-photon detectors (SNSPDs) was used to collect 10-minute resting-state data from 25 healthy adults and 5 patients with traumatic brain injury (TBI) diagnosed with DOC, including minimally conscious state (MCS) and coma, in the subacute phase. Photon arrival times were temporally gated to distinguish superficial and cortical-weighted tissue contributions. The blood-flow index (BFI) was extracted from gated autocorrelation functions, and LFOs were quantified using power spectral density within the Slow-5 (0.01-0.027 Hz), Slow-4 (0.027-0.073 Hz), and Slow-3 (0.073-0.198 Hz) bands. Compared to healthy controls, DOC patients exhibited altered resting-state LFO amplitude and spectral distribution, suggestive of altered neurovascular dynamics in severe brain injury. An auditory “smile” command was delivered to five healthy subjects, one MCS patient, and one unresponsive wakefulness syndrome (UWS) patient to assess task-evoked hemodynamic responses. During the task, healthy participants showed clear hemodynamic responses, whereas DOC patients demonstrated attenuated and more transient responses. Overall, TD-DCS provides a noninvasive, depth-resolved approach for assessing cerebral hemodynamics and residual cortical responsiveness, supporting its potential for bedside neurocritical-care monitoring.

## Introduction

Acute brain injury (ABI) is a leading cause of mortality and long-term disability, particularly among young adults ^1–4^ . While the primary injury occurs at the moment of trauma, secondary processes such as impaired autoregulation, ischemia, and inflammation can evolve over hours to days, driving further neurological deterioration ^5,6^. Continuous bedside monitoring of cerebral blood flow (CBF) is therefore highly desirable for detecting secondary injury and guiding timely intervention ^7–13^. Conventional imaging techniques such as magnetic resonance imaging (MRI), computed tomography (CT), and positron emission tomography (PET) provide detailed structural and functional information but are limited by their high cost, logistical complexity, and lack of feasibility for continuous bedside monitoring ^14^. Diffuse correlation spectroscopy (DCS) offers a non-invasive and portable alternative capable of tracking microvascular CBF in real time. Compared to near-infrared spectroscopy (NIRS), DCS provides greater sensitivity to cerebral blood flow and has shown promise in neurocritical care applications ^11,15,16^. However, standard continuous-wave (CW) DCS systems are limited in depth specificity and can be influenced by signals from superficial tissues such as the scalp and skull ^17–19^.

Time-domain DCS (TD-DCS), enhances depth sensitivity by isolating late-arriving photons that have traveled deeper into tissue ^20–22^. When implemented at longer wavelengths such as 1064 nm and combined with highly sensitive detectors such as SNSPDs, TD-DCS enables deeper penetration, improved signal-to-noise ratio, and higher permissible laser power, making it well-suited for adult human brain monitoring ^23–33^. While cerebral blood flow parameter provides one indication of brain health, spontaneous low-frequency oscillations (LFOs) in the range of 0.01-0.2 Hz are known to reflect important physiological processes including endothelial, neurogenic, and respiratory activity, and have been associated with cerebrovascular regulation and brain function ^31,34–53^. Optical bedside monitoring of cerebral hemodynamics in brain injury has been demonstrated using DCS (e.g.,^54–56,56–65^). However, few studies have focused on low-frequency oscillations (LFOs), and their depth-specific characterization in acute human brain injury remains largely unexplored.

Disorders of Consciousness (DOCs) represent a clinical spectrum of severe cerebral dysfunction, encompassing coma and chronic states of impaired consciousness. These chronic states include the Unresponsive Wakefulness Syndrome (UWS) (formerly known as the Vegetative State), characterized by preserved arousal with no evidence of purposeful behavior or awareness, and the Minimally Conscious State (MCS), defined by inconsistent but reproducible signs of awareness, such as visual tracking, localization, or command following ^66, 67^ .

While resting-state signals and oscillations can provide insight into baseline physiology, task-evoked functional responses may offer greater contrast for assessing consciousness states by detecting residual cortical reactivity. Auditory or command-driven paradigms such as familiar voices or simple motor tasks elicit reproducible hemodynamic responses in conscious patients but are absent in deeply unconscious states^45,68–70^ . These paradigms are commonly studied with functional MRI (fMRI), which is impractical in intensive care unit (ICU) settings. In this study, we incorporated auditory stimulation tasks, which elicit reproducible hemodynamic responses in patients with residual consciousness ^45–53,68^. In the present study, task-based auditory stimulation is included primarily as a qualitative illustration of depth-sensitive TD-DCS signal behavior, rather than as a primary marker of cognitive function or motor planning.

Time-domain diffuse correlation spectroscopy (TD-DCS) addresses these limitations by directly measuring cerebral blood flow (CBF) and its spontaneous fluctuations in a depth-sensitive, non-invasive manner. Unlike NIRS- or BOLD-based techniques that rely on surrogate markers of oxygenation, TD-DCS quantifies microvascular flow dynamics with high temporal resolution and deep-tissue sensitivity when implemented at 1064 nm with superconducting nanowire single-photon detectors. This configuration enables deeper tissue penetration, improved signal-to-noise ratio, and higher laser safety limits, making it a realistic alternative to fMRI in longitudinal consciousness assessment and neurocritical-care monitoring ^10,71–78^ .

By combining the portability and bedside practicality of fNIRS and EEG with the physiological specificity of perfusion imaging, TD-DCS offers a unique opportunity for repeated, real-time assessment of brain function in DOC patients without requiring transport or complex infrastructure. In this study we evaluate the utility of TD-DCS in both resting-state and task-evoked paradigms to detect residual cortical reactivity and differentiate levels of consciousness in critically ill patients as a potential bedside adjunct for neurocritical-care monitoring.

## Materials and Methods

### Study Design and Patient Details

This study is a retrospective analysis of a prospective observational protocol designed to evaluate cerebral blood flow (CBF) in healthy individuals and patients with traumatic brain injury (TBI). A total of 25 healthy adults (14 males, 11 females; mean age 26.9 ± 4.8 years) were recruited for resting-state measurements, and an additional cohort of five healthy subjects (5 males; mean age 29.0 ± 5.5 years) participated in an auditory “smile” task to assess task-evoked hemodynamic responses. The auditory “smile” task consisted of repeated verbal commands (“smile”) delivered at fixed intervals to elicit cortical hemodynamic activation, as described below.

Six TBI patients were enrolled between 2022 and 2025 from the University of Cincinnati Medical Center and Stony Brook University Hospital. Of these, five patients (two diagnosed with minimally conscious state [MCS] and three with coma) underwent resting-state TD-DCS measurements. One additional patient diagnosed with unresponsive wakefulness syndrome (UWS) underwent task-based TD-DCS measurements only, using a bilateral probe configuration. A summary of subject diagnoses, clinical characteristics, and experimental inclusion is provided in Supplementary Table S1. For resting-state analyses, patients diagnosed with MCS (n = 2) and coma (n = 3) were analyzed together as a DOC cohort due to the limited sample size of individual diagnostic subgroups. Task-based measurements in DOC patients are presented descriptively at the individual-subject level.

Resting-state recordings consisted of a single 10-minute acquisition performed in the intensive care unit (ICU) during routine clinical care. Task-based recordings consisted of an auditory “smile” command delivered approximately every 60 seconds for a total of five repetitions ^79–81^. No experimental interventions were introduced, and no restrictions were placed on medical treatment during the data acquisition period. All healthy participants provided written informed consent under protocols approved by the Institutional Review Boards (IRBs) of Wright State University and Stony Brook University. The study was conducted in accordance with the Declaration of Helsinki and approved by the IRBs of Wright State University, Stony Brook University, and the University of Cincinnati Medical Center. For TBI patients, informed consent was obtained from legally authorized representatives in accordance with these IRB approvals.

### TD-DCS System for Data Acquisition

The TD-DCS method has been employed in numerous previous studies, demonstrating its feasibility for clinical use ^82–91^ . Figure 1a-c illustrates the components of the system and probe placement. The optical imaging probe was positioned on the right forehead, between EEG scalp locations F4 and F8, to ensure consistent and anatomically relevant placement. The probe incorporated a 600-µm-core multimode source fiber (NA = 0.39, FT600EMT, Thorlabs, NJ, USA) and an 8.2-µm-core detector fiber (SMF-28, Thorlabs, NJ, USA), which is few-mode at 1064 nm^92^. The source-detector separation (SD) was fixed at 15 mm, a deliberate choice reflecting the established TD-DCS trade-off between cortical-weighted sensitivity and photon statistics, in which depth sensitivity is governed primarily by photon time-of-flight and further enhanced through late temporal gating rather than separation alone ^17,18,21,23^. All fibers were aligned through a 5 mm right-angled prism and embedded in a custom-designed, flexible 3D-printed housing optimized for ergonomic use in clinical environments. The probe’s contact footprint was approximately 25 mm × 25 mm, slightly larger than a US quarter, minimizing scalp contact while enabling secure and reproducible placement. The assembly was affixed using medical-grade Tegaderm film (3M, Saint Paul, MN) following sterilization with alcohol swabs. Reflected photons at 1064 nm were detected using four superconducting nanowire single-photon detectors (SNSPDs), which were interfaced with a 4-channel time-correlated single-photon counting (TCSPC) system (HydraHarp400, PicoQuant GmbH, Berlin, Germany) ^93,94^. During the initial phase of the study, the healthy resting-state cohort (H1-H25) and one comatose TBI patient (Coma-1) measured at the University of Cincinnati Medical Center, a 1064 nm pulsed seed laser (QC2D106P-64D0, QDLaser Inc., Kanagawa, Japan) operating at an 80 MHz repetition rate with a 250 ps pulse width was used. For all remaining subjects, including patients with disorders of consciousness, as well as for all functional task (“smile”) measurements, a 1064 nm pulsed laser (VisIR-1064-HC High-Coherence, PicoQuant GmbH, Berlin, Germany) operating at an 80 MHz repetition rate with a 400 ps pulse width was used. The beam exiting the prism face was expanded to approximately 4.5 mm in diameter using a diffuser film to ensure uniform illumination across a large beam area. The average power was kept below ANSI safety limits, and each detection channel operated at a photon count rate of ∼1 × 10⁷ counts per second (cps).

**Figure 1.**
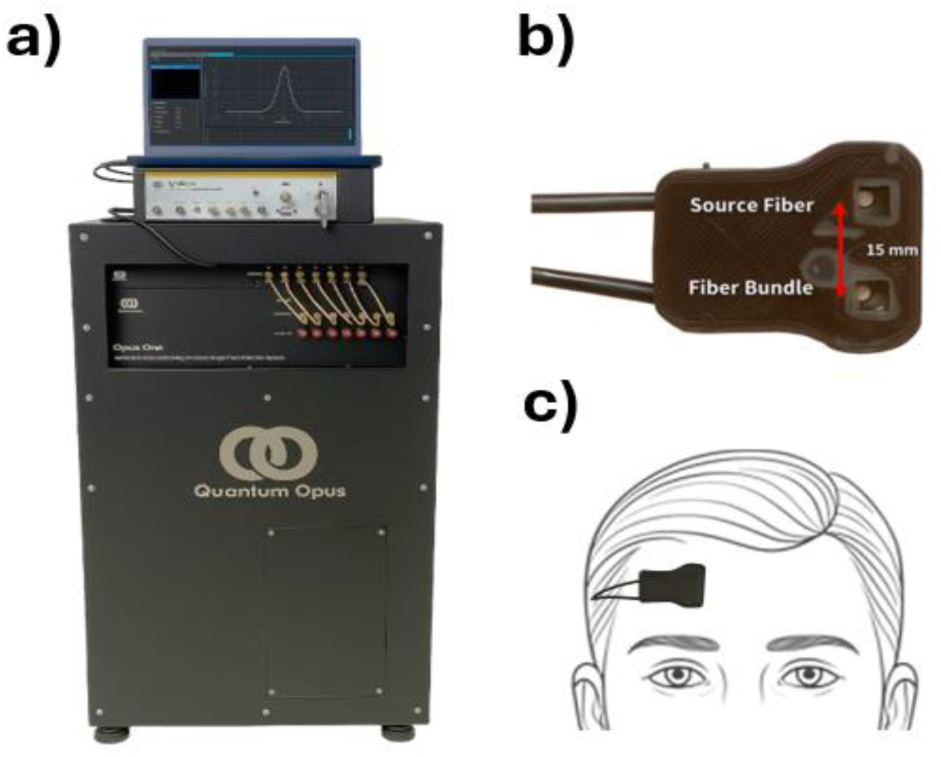
Overview of the time-domain diffuse correlation spectroscopy (TD-DCS) system. a) Quantum Opus superconducting nanowire single-photon detector (SNSPD) system with the VisIR pulsed laser (1064 nm) mounted on top. The display shows the photon distribution of time-of-flight (DTOF). b) Close-up view of the custom 3D-printed probe with a 15 mm source-detector separation between the source and detector fibers. c) Example of the probe placed on the right forehead of a human subject for cerebral blood flow measurements.

### Data Analysis

The analyses below were designed to test protocol-specific hypotheses regarding depth-resolved resting-state dynamics and task-evoked depth-dependent hemodynamic signals.

### Resting-State Analysis

Time-tagged photon arrival data were recorded continuously. Resting-state measurements consisted of a single 10-minute acquisition per subject. Data processing was performed using a custom MATLAB routine on a high-memory computing node with 1.5 TB of RAM. The instrument response function (IRF) was characterized by placing a thin scattering layer (Teflon tape, 3M) between the source and detector fibers during a 30-second calibration, resulting in a full width at half maximum (FWHM) of ∼400 ps for the VisIR-1064-HC ^95^ . Two temporal gates were selected based on the distribution of time-of-flight (DTOF). The early gate (EG), centered at 0.20 ns with a width of 100 ps, was positioned just after the DTOF peak to emphasize photons that predominantly sampled superficial tissue layers (Fig. 2a). The late gate (LG), centered at 1.03 ns with a width of 250 ps, was placed on the decaying slope of the DTOF to enhance sensitivity to deeper tissue regions (Fig. 2d). Figure 2 illustrates these gating definitions using data from a representative comatose patient. Gate positions were defined relative to the peak of the combined DTOF across all channels to maintain consistent depth sensitivity among subjects. This strategy was designed to capture photons from deeper tissue layers while minimizing contamination from superficial dynamics. It aligns with prior findings that optimally selected narrow time gating (e.g., 100 ps) with sufficient SNR can enhance depth specificity in TD-DCS ^96^. As motivated by Tagliabue *et al* ^97^, who demonstrated spatial and temporal optical heterogeneity in injured brains, we performed subject-specific fitting of the temporal point spread function (TPSF) to extract the absorption (𝜇_𝑎_) and reduced scattering 𝜇′_*s*_ coefficients, which were then used as fixed *a priori* inputs for the analytical intensity autocorrelation function (g_2_) fitting to quantify the blood flow index (BFI). The TPSF fitting was performed using an analytical solution of the time-domain diffusion equation in reflectance geometry assuming a semi-infinite homogeneous medium with extrapolated boundary conditions ^98,99^.

**Figure 2.**
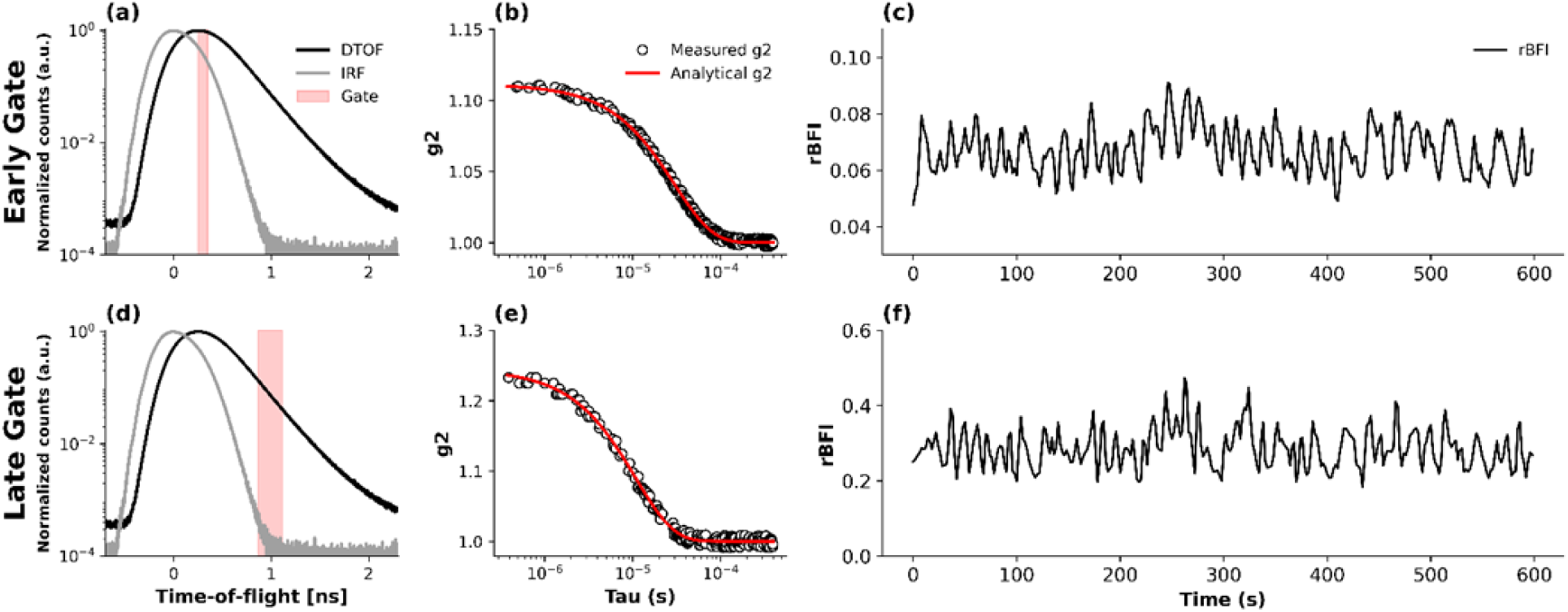
Early- and late-gate TD-DCS analysis for a representative comatose patient. (a,d) Normalized distributions of time-of-flight (DTOF, black) and instrument response function (IRF, gray), with the selected gate window shown in red for the early and late gates, respectively. (b,e) Corresponding normalized intensity autocorrelation curves g_2_(τ) (black circles) and analytical fits (red lines) used to estimate the blood-flow index (BFI). (c,f) Temporal evolution of the relative blood-flow index (rBFI) derived from the early and late gates, respectively, showing increased fluctuation amplitude and deeper sensitivity for the late-gate signal.

Under this modeling framework, the extracted optical properties should be interpreted as effective bulk parameters that reflect the weighted contribution of all tissues sampled by the photon time-of-flight distribution, rather than depth-resolved or layer-specific tissue properties. This approach accounts for inter-subject variability in optical properties, which is particularly relevant in TBI patients where extracerebral tissue characteristics may vary substantially. Representative TPSF and IRF fits for healthy, MCS, and comatose subjects are provided in Supplementary Fig. S1 to illustrate fitting quality; optical property values are shown for completeness and are not interpreted or compared across diagnostic groups. The g_2_ functions were extracted directly from each temporal gate using delay times ranging from 5 × 10^−7^s to 1 × 10^−3^ s. Autocorrelations were calculated using 5 s integration windows with 1 s step size, producing overlapping data segments to enhance temporal resolution (Figs. 2b, e). The resulting g_2_ curves were fitted to an analytical model derived from the time-domain diffusion equation assuming a semi-infinite homogeneous medium geometry ^17,20,100^. Although this geometry does not explicitly model layered head anatomy or depth-resolved photon trajectories, depth sensitivity was achieved experimentally through time-of-flight–based temporal gating, with the late gate providing increased sensitivity to deeper cerebral tissue and reduced sensitivity to extracerebral contributions. Because detection was performed with an 8.2 µm-core few-mode fiber, the measured coherence factor (β) values were lower than those typically obtained in single-mode DCS systems (β ≈ 0.5), resulting in smaller g_2_ amplitudes (Figs. 2b, e). Although this configuration reduces temporal coherence, it substantially improves the signal-to-noise ratio (SNR), which is critical for reliable estimation of cerebral blood flow in human measurements. Final relative blood-flow index (rBFI) values were obtained by fitting g_2_ curves averaged across all four detection channels (Figs. 2c, f). For resting-state measurements, absolute BFI time series were used to compute mean flow values and low-frequency oscillation (LFO) metrics.

### Spectral Analysis of Low-Frequency Oscillations (LFOs)

In addition to evaluating cerebral blood flow changes, we analyzed LFOs in the blood flow index as potential indicators of cerebrovascular function. LFOs were defined as spontaneous fluctuations occurring below 0.5 Hz and were examined within three descriptive frequency bands (Slow-5, Slow-4, and Slow-3), as summarized in Table 1. This frequency-band nomenclature follows prior resting-state fMRI and optical neuroimaging literature ^34,48,101–106^, where low-frequency oscillations are commonly subdivided into Slow-5, Slow-4, and Slow-3 bands for spectral characterization^101–103^. Consistent with common fMRI and fNIRS practice, the broader low-frequency range (≈0.01-0.1 Hz) is generally interpreted as reflecting neurovascular coupling in aggregate; the subdivision into Slow-5, Slow-4, and Slow-3 bands is used here to facilitate comparative, depth-resolved spectral analysis rather than to imply distinct or exclusive physiological generators. Bands I and II are listed in Table 1 for completeness but were not analyzed or interpreted separately in this study; band-specific comparisons focus on Slow-5, Slow-4, and Slow-3 (Bands V-III).

**Table 1:** Frequency bands for low-frequency oscillations (LFOs) with respect to band-1 to band-5, and their associated descriptor.

| Band | Frequency Range (Hz) | Descriptor |
| --- | --- | --- |
| V | 0.01-0.027 | Very-low-frequency LFO |
| IV | 0.027-0.073 | Low-frequency LFO |
| III | 0.073-0.198 | Higher low-frequency LFO |
| II | 0.198-0.5 | Higher-frequency physiological range / respiration contaminated |
| I | 0.5-0.75 | Higher-frequency physiological range / noise |

Prior to spectral analysis, the resting-state BFI time series were detrended using a second-order polynomial to remove slow baseline drifts. The signals were then band-pass filtered between 0.01 and 0.4 Hz using a zero-phase Butterworth filter (2nd order) to isolate low-frequency oscillations while avoiding phase distortion. Power spectral density (PSD) was computed using the fast Fourier transform (FFT) applied to the full 10-minute resting-state BFI time series for each subject. The total amplitude of low-frequency oscillations was quantified using the amplitude of low-frequency fluctuations (ALFF), defined as the square root of the integrated PSD over the 0.01-0.4 Hz range. To compare spectral shapes across subjects, each PSD was normalized by its own total low-frequency power and interpolated onto a common frequency axis between 0.01 and 0.4 Hz (see Figs. 3c, g for representative group-averaged normalized PSDs). A sensitivity analysis examining the effect of the PSD normalization bandwidth on normalized spectral estimates is provided in Supplementary Fig. S2. Because PSD.n and LFO.n are normalized by the total 0.01-0.4 Hz power, these metrics describe spectral redistribution (band reorganization) rather than absolute oscillation amplitude.

**Figure 3.**
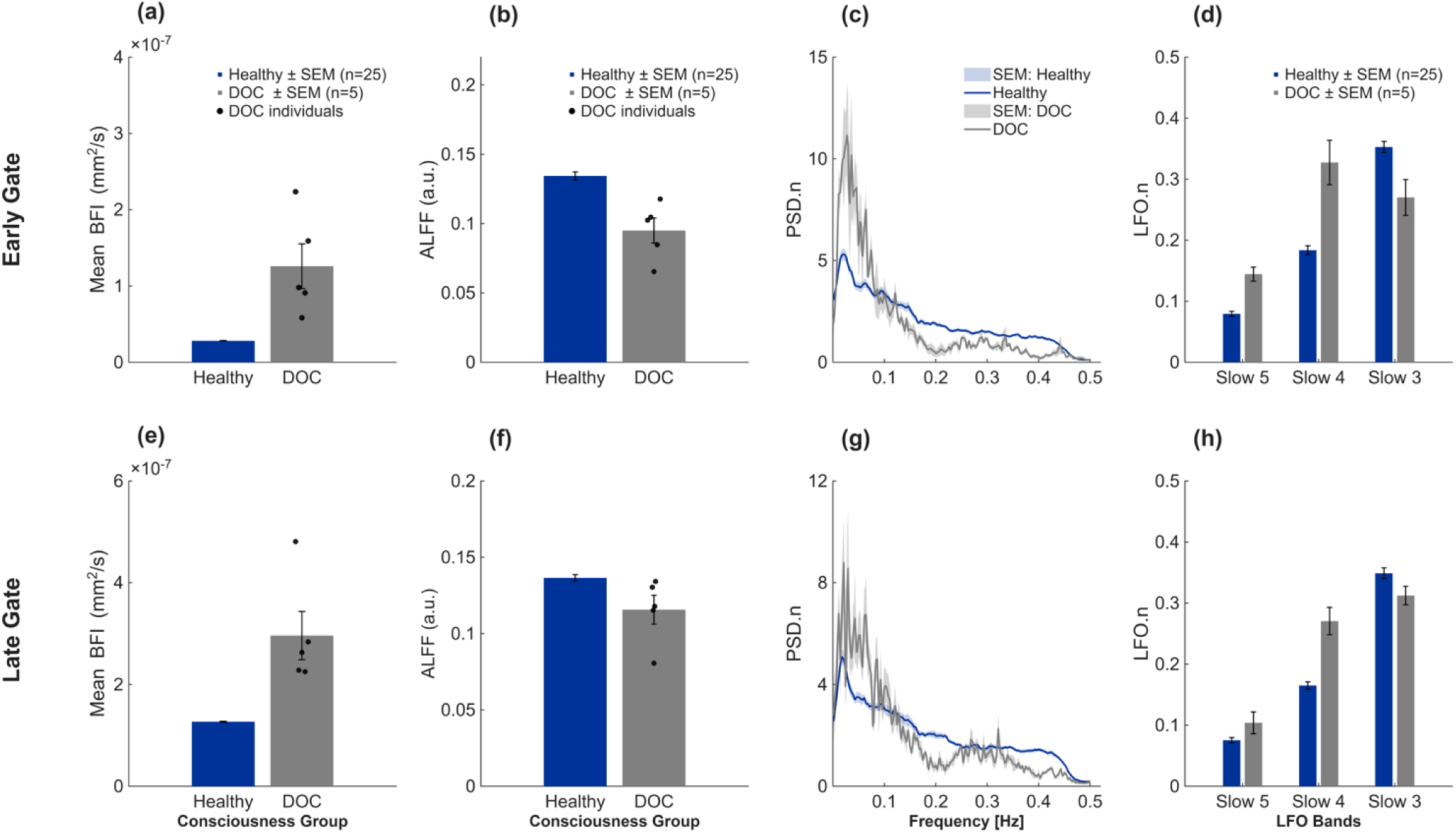
Depth-resolved resting-state cerebral blood flow and low-frequency oscillations in healthy controls and patients with disorders of consciousness. (a-d) Early-gate (EG) results, preferentially weighted toward superficial tissues. (e-h) Late-gate (LG) results, providing enhanced sensitivity to deeper (cortical-weighted) signals. (a,e) Mean absolute blood-flow index (BFI, mm²/s). (b,f) Amplitude of low-frequency fluctuations (ALFF; 0.01-0.4 Hz). (c,g) Normalized power spectral density (PSD.n) of the resting-state BFI signal (mean ± SEM). (d,h) Normalized band-limited LFO power (LFO.n), defined as the fraction of total 0.01-0.4 Hz power contained within Slow-5 (0.01-0.027 Hz), Slow-4 (0.027-0.073 Hz), and Slow-3 (0.073-0.198 Hz). Bars indicate group mean ± SEM for healthy controls (n = 25) and DOC patients (n = 5; minimally conscious state and coma); black markers denote individual DOC subjects in panels (a,b,e,f). Results are presented descriptively due to the limited DOC sample size.

Band-limited power in each LFO range (𝑃_𝐵_) was obtained by integrating the unnormalized PSD over the corresponding frequency interval. Normalized band power was defined as 𝑅_𝐵_ = 𝑃_𝐵_⁄𝑃_total_, where 𝑃_total_ is the total integrated PSD over 0.01-0.4 Hz; thus 𝑅_𝐵_ represents the fraction of total 0.01-0.4 Hz BFI power allocated to Slow-5, Slow-4, or Slow-3.

Mean and standard error of 𝑃_total_ and 𝑅_𝐵_ were calculated for both the healthy control group and the DOC cohort (n = 5). The summary statistics are reported for descriptive purposes only, and individual subject values are explicitly visualized to reflect inter-subject variability in the context of the limited sample size.

We note that arterial blood pressure (ABP) was not recorded in this feasibility study. Unlike transfer-function analyses of autoregulation that require ABP as an input signal (e.g., ^107–112^), our approach focuses on the output fluctuations of the TD-DCS blood-flow index and on the relative distribution of power across physiologically motivated frequency bands.

### Task-Based (Auditory “Smile”) Measurements

Task-based recordings consisted of an auditory “smile” command delivered approximately every 60 s for a total of five repetitions, resulting in an acquisition duration of approximately 5 min per subject. For task-based analyses, relative blood-flow index (rBFI), defined as BFI normalized to a pre-stimulus baseline, was used to characterize stimulus-evoked hemodynamic responses. Task-based analyses were performed to explore whether stimulus-evoked cortical hemodynamic responses observed in healthy subjects could also be detected at the individual-subject level in patients with disorders of consciousness, including MCS and UWS.

Early-gate and late-gate rBFI time series were detrended and band-pass filtered to suppress slow baseline drift and high-frequency noise while preserving task-relevant dynamics. Expected stimulus onset times were defined based on the experimental protocol, and peri-stimulus windows were extracted for each trial. Local maxima in EG and LG signals were identified within predefined temporal search windows following stimulus onset to account for inter-subject variability in response timing. Extracted trials were baseline-corrected using a pre-peak interval and averaged within each subject.

Because late-gate TD-DCS measurements are inherently low signal-to-noise due to reduced photon counts at longer time-of-flight, particularly in clinical bedside measurements, a state-space modeling approach was employed to robustly infer the latent cortical hemodynamic response. The observed LG rBFI signal was modeled as a combination of a latent task-evoked response and a superficial contribution proportional to the EG signal. The latent response was represented using a set of Gaussian basis functions convolved with the stimulus time course. A Kalman filter with Rauch-Tung-Striebel smoothing was applied to estimate the underlying latent state by combining temporal continuity constraints with measurement uncertainty ^113,114^. In this formulation, the latent state comprised task-evoked hemodynamic response weights and a time-varying superficial coupling coefficient, while the observation model related these latent variables to the measured late-gate rBFI signal. The inferred LG signal, with superficial contributions regressed out to reduce extracerebral contamination, was used for subsequent block averaging and descriptive comparison across subjects.

## Results

### Resting-State and Task-Based TD-DCS Measurements

#### Resting-State Cerebral Blood Flow and LFOs: Healthy vs DOC

Resting-state TD-DCS measurements were obtained from healthy controls (n = 25) and from patients with disorders of consciousness (DOC; n = 5), including individuals diagnosed with minimally conscious state and coma. Only subjects who completed resting-state recordings were included in this analysis; task-only subjects were excluded. In particular, the post-surgical unresponsive wakefulness syndrome (UWS) patient who participated only in the auditory task was not included in the resting-state analysis.

Depth sensitivity was achieved using time-of-flight-based temporal gating, with the early gate (EG) preferentially sampling superficial tissues and the late gate (LG) providing enhanced sensitivity to deeper, cortical-weighted signals **(Fig. 3a, e)**. Mean blood-flow index (BFI), used here as a proxy for cerebral blood flow, was summarized descriptively for both groups, with individual subject values explicitly shown for the DOC cohort. In the superficial compartment (EG; **Fig. 3a)**, DOC patients exhibited descriptively higher BFI values compared to healthy controls, accompanied by substantial inter-subject variability. A similar pattern was observed in the cortical-weighted compartment **(LG; Fig. 3e),** where DOC patients again demonstrated elevated BFI relative to healthy participants. These observations suggest altered baseline perfusion dynamics in disorders of consciousness, though no statistical inference is made due to the limited patient sample size. Low-frequency oscillations (LFOs) in the resting-state BFI signal were further examined using the amplitude of low-frequency fluctuations (ALFF; 0.01-0.4 Hz) to assess depth-dependent oscillatory dynamics. In the superficial compartment **(EG; Fig. 3b),** healthy participants exhibited descriptively higher ALFF values compared to the DOC cohort, whereas in the cortical-weighted compartment **(LG; Fig. 3f),** group differences were less pronounced, with overlapping values observed across subjects.

Normalized power spectral density analyses (PSD.n; **Fig. 3c, g)** revealed systematic depth-dependent spectral reorganization. Healthy controls exhibited a broader distribution of power across the low-frequency range, while DOC patients showed a relative shift toward very low frequencies (< 0.1 Hz), particularly in cortical-weighted late-gate measurements. This pattern indicates a greater concentration of spectral power at frequencies below 0.1 Hz in the DOC cohort, particularly in late-gated measurements. Normalized band-limited LFO metrics (LFO.n; Fig. 3d,h) further quantified this spectral distribution. Healthy participants demonstrated relatively greater contributions from higher-frequency LFO components, whereas DOC patients exhibited increased dominance of slower oscillatory bands. Together, these findings show descriptively different baseline blood-flow magnitude and low-frequency oscillatory structure between healthy controls and the DOC cohort, with depth-dependent differences observed across early- and late-gated signals.

#### Illustrative Depth-Sensitive Hemodynamic Signals During Auditory Stimulation

To qualitatively assess depth-dependent hemodynamic sensitivity during a task paradigm, we analyzed early- and late-gated TD-DCS responses during an auditory stimulation protocol in healthy controls (n = 5) and a patient diagnosed with unresponsive wakefulness syndrome (UWS; n = 1). Early- and late-gated measurements were interpreted as being differentially weighted toward superficial extracerebral and deeper cerebral tissue compartments, respectively, consistent with prior time-domain optical studies exploiting photon time-of-flight information and depth-sensitive observables to reduce superficial contamination ^115,116^. Task-locked responses were estimated using a Kalman-based regression framework with the stimulation paradigm as a regressor, followed by trial-wise baseline correction and block averaging.

#### Early-Gated Signals

In healthy subjects, early gated signals exhibited large, task-locked increases in relative blood flow index (rBFI), with peak amplitudes occurring during the stimulation window (Fig. 4.a). These responses were robust across trials and substantially larger than corresponding late-gated signals. Given the strong sensitivity of early-arriving photons to superficial tissue layers, these pronounced task-locked EG responses are consistent with extracerebral physiological contributions, including scalp and autonomic vascular responses, as widely reported in time-domain fNIRS and diffuse optical studies^115–117^. In contrast, the UWS patient demonstrated minimal modulation in early-gated rBFI during auditory stimulation, with signals remaining near baseline throughout the task period. This absence of early-gated task reactivity is consistent with reduced superficial physiological engagement during stimulation and further highlights the sensitivity of EG TD-DCS measurements to superficial rather than cerebral hemodynamic changes.

**Figure 4.**
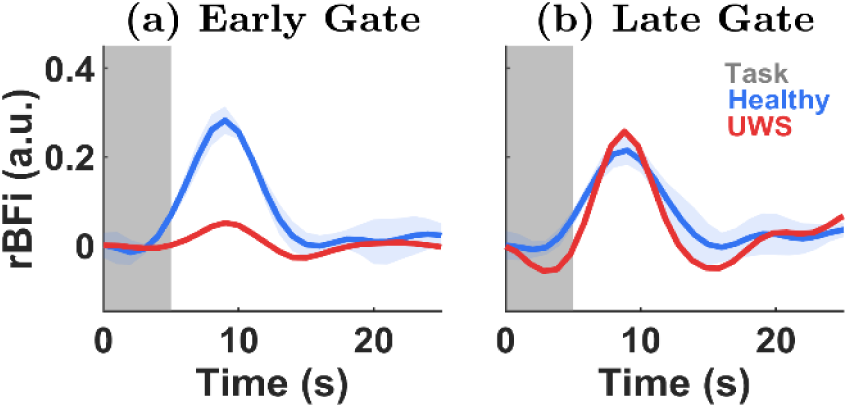
Depth-sensitive TD-DCS responses during a 5-s auditory stimulation task. Early gated (a) and late gated (b) relative blood flow index (rBFI) responses in healthy controls (blue, mean ± SEM) and a UWS patient (red). Shaded region indicates the 5-s task period.

#### Late-Gated Signals

Late-gated (LG) signals exhibited smaller-amplitude but structured task-locked dynamics in both healthy subjects and the UWS patient (Fig. 4. b). Compared to early-gated measurements, late-gated responses were temporally smoother and showed reduced variability, consistent with increased weighting toward deeper tissue compartments and reduced sensitivity to superficial physiology. This behavior closely parallels prior time-domain optical findings demonstrating that depth-weighted or late-photon observables substantially suppress task-locked superficial contributions present in attenuation-based or early-weighted measurements ^115,118^.

Notably, the UWS patient exhibited late-gated response amplitudes comparable to, and in some instances exceeding, those observed in healthy controls. Similar increases in hemodynamic response amplitude in UWS relative to healthy subjects have been reported in prior neuroimaging studies and interpreted as reflecting altered or dysregulated neurovascular coupling rather than preserved cognitive processing^119^. Accordingly, these late-gated task responses are presented descriptively and interpreted cautiously.

## Discussion

Accurate differentiation between UWS and MCS remains difficult, with misdiagnosis rates of 30–40% when relying solely on clinical examination ^45,120–122^. Neuroimaging and neurophysiological modalities, including fMRI ^79,123,124^, PET ^125–128^, EEG ^126,126,129,129,130,130–133^, and NIRS ^80,134–136^, have enabled detection of MCS/covert consciousness, in which patients demonstrate preserved cortical activation despite absent behavioral output. However, these techniques are limited by either lack of portability (fMRI, PET) or depth specificity (EEG, NIRS), constraining their feasibility for repeated bedside assessments in critically ill patients. Electroencephalography (EEG) provides bedside compatibility and high temporal resolution but suffers from low spatial specificity, susceptibility to artifacts, and reduced sensitivity to deep cortical responses, particularly in patients with skull defects ^132,137,138^. Functional NIRS has recently shown promise in detecting task-evoked responses using personalized motor-speech imagery and covariance-based classifiers ^139–141^, yet it remains limited by superficial contamination and reliance on surrogate oxygenation measures. In this context, there remains a need for a portable, depth-sensitive method capable of assessing cerebral physiology at the bedside.

This study demonstrates a novel application of TD-DCS for depth-sensitive assessment of CBF and spontaneous LFOs in healthy individuals and patients with TBI diagnosed with disorders of consciousness (DOC). Using a portable 1064 nm TD-DCS system with SNSPDs, we obtained depth-specific hemodynamic signatures that distinguish superficial and cortical-weighted vascular dynamics ^26,27,142^. Given the limited sample size of individual diagnostic subgroups, resting-state findings are interpreted at the level of healthy controls versus a combined DOC cohort, and observations within DOC patients are treated as descriptive rather than inferential.

Compared to healthy controls, DOC patients exhibited altered resting-state LFO characteristics in both early- and late-gated signals, with larger shifts in the early gate and attenuated but directionally consistent redistribution trends in the late gate. In our cohort, ALFF tended to be lower in DOC patients in both gates, while normalized spectral analyses demonstrated increased relative weighting of very low frequencies (<0.1 Hz) and greater Slow-5/Slow-4 fractions with reduced Slow-3 contribution. These observations are broadly consistent with prior fNIRS and fMRI studies reporting disrupted resting-state hemodynamic oscillations and altered neurovascular dynamics in severe brain injury and disorders of consciousness (e.g. ^80,143–145^). In particular, optical and fMRI studies such as Schulthess et al.^81^ and Kazazian et al.^80^ have reported reduced spontaneous or task-evoked low-frequency activity in unresponsive wakefulness syndrome, supporting the interpretation that preserved LFO may reflect residual neurovascular reactivity and cortical viability. These findings contrast with acute-phase animal studies (e.g., White et al.^146^), which reported increased LFOs shortly after injury, potentially reflecting metabolic instability and/or pathological vasomotion during the hyperacute phase. In the subacute clinical setting of the present study, the observed spectral slowing and redistribution of LFO content may be more consistent with dysregulated vasomotor control and altered vasomotor control and altered neurovascular regulation rather than acute hyperacute increases or inflammation. While DOC patients demonstrated heterogeneous resting-state profiles, no statistical separation between minimally conscious and comatose patients is inferred from the present data due to the limited subgroup sizes. Instead, the observed inter-subject variability highlights the complexity of cerebrovascular dysfunction following severe brain injury and underscores the need for larger cohorts to determine whether depth-resolved spectral metrics can reliably differentiate diagnostic categories or track recovery trajectories.

Mean BFI was higher in late-gated signals than in early-gated measurements, particularly in DOC patients. Low-frequency spectral features were observed in both gates, with larger shifts in the early gate and attenuated but directionally consistent redistribution trends in the late gate. The observed elevation in baseline BFI in DOC patients is interpreted cautiously and is not attributed to preserved neural activity. Instead, increased BFI may reflect altered cerebrovascular tone and dysregulated neurovascular control after severe brain injury, including impaired autoregulatory mechanisms and disrupted flow-metabolism coupling, as well as systemic influences (e.g., ventilation/PaCO_2_, sedation, vasoactive medications). Similar elevations in baseline CBF or perfusion have been reported in subacute TBI and disorders of consciousness and are often interpreted as reflecting dysregulated cerebrovascular control rather than task-evoked functional activation ^11,32,147^.

In contrast, normalized low-frequency spectral metrics (PSD.n and LFO.n) emphasize how the remaining fluctuations are distributed across frequencies and therefore support interpretation in terms of spectral reorganization. Such alterations in low-frequency spectral content have been associated with altered autoregulation and vasomotor dysregulation in prior optical studies ^38,41,43,45,50,51,53,148^. Both early and late gates showed depth-dependent spectral redistribution across LFO bands, with larger shifts in the early gate and similar but attenuated trends in the late gate. The persistence of these trends in the late gate is consistent with a deeper-weighted contribution and highlights the utility of time-gating for separating superficial-weighted and cortical-weighted dynamics in neurocritical-care applications. In addition to resting-state analysis, we evaluated functional reactivity using a brief auditory “smile” protocol. The task-based TD-DCS results further illustrate the differential sensitivity of early- and late-gated measurements to superficial versus depth-weighted hemodynamic contributions. In healthy subjects, large task-locked responses observed in the early gate are consistent with the strong sensitivity of early-arriving photons to superficial extracerebral physiology, which may include autonomic or arousal and systemic coupling effects during task engagement. The absence of comparable early-gated modulation in the UWS patient suggests the dependence of early-gated signals on superficial physiological engagement rather than cerebral hemodynamics. In contrast, late-gated signals exhibited lower amplitude and smoother temporal dynamics across both groups, consistent with increased weighting toward deeper tissue compartments and partial suppression of superficial contributions. The presence of structured late-gated responses in the UWS patient, with amplitudes comparable to or exceeding those observed in healthy controls, is consistent with prior neuroimaging studies reporting altered hemodynamic responses in disorders of consciousness ^80,149^. Such responses have been interpreted as reflecting dysregulated neurovascular coupling or altered cerebrovascular dynamics rather than preserved cognitive or motor processing. Importantly, these task-based findings are presented descriptively and are not intended to infer localized cortical activation, motor planning, or functional network engagement. Instead, they serve to qualitatively demonstrate how photon time-of-flight gating alters task-locked signal behavior in TD-DCS measurements in brain-injured patients. Together with the resting-state LFO analyses, these results emphasize the importance of depth-sensitive approaches for interpreting optical hemodynamic signals in both healthy and severely brain-injured populations.

Together, these results highlight the potential of TD-DCS to provide real-time, depth-resolved assessment of cerebral hemodynamic dynamics and neurovascular integrity in critically ill patients at the bedside. The ability to non-invasively measure cerebral LFOs and task- or state-related BFI fluctuations at the bedside has significant clinical implications. Current neuro-monitoring tools in the ICU are either invasive (e.g., intracranial pressure monitors), lack depth sensitivity (e.g., NIRS), or are not practical for continuous use (e.g., MRI, PET). TD-DCS bridges this gap by offering portable, real-time monitoring of both baseline perfusion and depth-weighted hemodynamic signals. In neurocritical care, LFO patterns may serve as indicators of cerebrovascular health, injury severity, and recovery potential ^42,45,51^. Longitudinal tracking of LFOs and hemodynamic responses could help clinicians identify patients at risk of secondary injury, evaluate the effectiveness of therapeutic interventions, and guide decisions around sedation, rehabilitation, and prognosis.

While our findings are promising, several limitations warrant consideration. The patient data are limited to five TBI cases, which restricts generalizability. Given the small MCS sample size (n = 2), group-level resting-state findings should be interpreted as preliminary and descriptive; larger cohorts will be required to assess the impact of focal pathologies such as hemorrhage on depth-resolved hemodynamic metrics. Larger cohorts are needed to validate LFO spectral changes as reliable markers of injury severity and recovery. Although time-gating improves cerebral sensitivity, residual contamination from superficial tissues cannot be fully ruled out. Similarly, optical properties (𝜇_*a*_, 𝜇′_*s*_) estimated from time-resolved TPSF fitting using a homogeneous semi-infinite diffusion model should be interpreted as effective bulk parameters reflecting the weighted contribution of both superficial and cerebral tissues. In patients with prior cranial surgery, such as the MCS case included here, extracerebral structural alterations may influence the fitted 𝜇′_*s*_ values and should not be interpreted as direct markers of cortical microstructure ^150,151^. The β values derived from our g_2_ fits were lower than standard CW-DCS systems due to the use of few-mode fibers, although it enabled sufficient photon throughput for feasibility of human brain measurements. Prior optical studies used ABP as an input function to assess autoregulation through transfer-function analysis ^108,110,148,152–156^ . In this feasibility study, ABP was not recorded, so our interpretation relied on the output signal of TD-DCS. By normalizing PSD to total power, we focused on the relative distribution of oscillatory activity across bands. While this approach does not capture input-output coupling, it provides complementary information on depth-dependent cerebrovascular dynamics. Future work will incorporate systemic inputs for a more complete assessment of autoregulation. Combining TD-DCS with other modalities such as EEG or fNIRS may enhance spatial resolution and signal interpretation ^38,111,112,157–159^ . Future hardware development could explore optimized geometries that maintain high count rates while covering more spatial coverage. Longitudinal studies will be important to test whether TD-DCS metrics track recovery or deterioration over time.

## Conclusions

This study demonstrates the utility of time-domain diffuse correlation spectroscopy (TD-DCS) as a non-invasive, depth-sensitive approach for monitoring cerebral blood flow (CBF) and spontaneous low-frequency oscillations (LFOs) in the human brain. Resting-state analyses revealed altered depth-resolved LFO characteristics in patients with acute brain injury and disorders of consciousness compared with healthy controls, with larger shifts in early-gated signals and attenuated but directionally consistent redistribution trends in late-gated signals. These findings are suggestive of altered cerebrovascular regulation following severe brain injury. By exploiting photon time-of-flight information, TD-DCS enabled separation of superficial and deeper hemodynamic contributions, highlighting the importance of depth-sensitive measurements for accurate interpretation of optical signals in neurocritical care. Task-based measurements were included as a qualitative illustration of depth sensitivity and demonstrated distinct early- and late-gated signal behavior, without implying localized cortical activation or preserved cognitive function. Together, these results underscore the potential of TD-DCS as a portable bedside tool for depth-resolved assessment of cerebral hemodynamics and vascular dynamics in critically ill patients. With further technical refinement and validation in larger cohorts, TD-DCS may complement existing neuro-monitoring modalities by enabling longitudinal, real-time tracking of cerebrovascular function and recovery trajectories in traumatic brain injury.

## Code and Data Availability

Data may be provided by the corresponding author upon reasonable request.

## Data Availability

All data produced in the present study are available upon reasonable request to the authors

## Acknowledgements

The authors acknowledge the funding support from NIH R01 (NIBIB Brain Initiative (7R01EB031759-03). We thank Nick Bertone (Picoquant Inc.) for providing the demo unit of HydraHarp400 for the time domain acquisition system. A pre-print version of this manuscript can be found on medRxiv, https://doi.org/10.1101/2025.08.31.25334647. ChatGPT (GPT–4) was used to assist with grammar and language editing during the preparation of this manuscript.

---

**Supplementary Table S1.** Clinical characteristics and experimental inclusion of all subjects.

| Subject | Diagnosis | Age | Sex | Hemorrhage | Surgery | Resting-State<br>(Fig. 3) | Smile Task<br>(Fig. 4) | Notes |
| --- | --- | --- | --- | --- | --- | --- | --- | --- |
| H1–H25 | Healthy | 21–39 (mean<br>26.9 ± 4.8) | 14 M / 11 F | No | No | Yes | No | Resting-state cohort |
| H26–H30 | Healthy | 25–35 (mean<br>29.0 ± 5.5) | 5 M | No | No | No | Yes | Smile-task cohort |
| MCS-1 | MCS | 40 | M | No | No | Yes | Yes | Resting-state only |
| MCS-2 | MCS | 75 | M | Yes<br>(left temporal) | No | Yes | No | Resting-state only |
| UWS-1 | UWS | 40 | M | Yes<br>(multicompartmental) | Yes<br>(decompressive craniectomy + skull reattachment) | No | Yes | Dual-probe bilateral + task |
| Coma-1 | Coma | 40 | M | No | No | Yes | No | Resting-state only |
| Coma-2 | Coma | 40 | M | No | No | Yes | No | Resting-state only |
| Coma-3 | Coma | 70 | M | No | No | Yes | No | Resting-state only |

